# General practitioners ending their practice and impact on patients’ health, health care use and mortality. A protocol for national registry cohort studies in Norway, 2006 to 2021

**DOI:** 10.1101/2023.01.25.23284995

**Authors:** Kristin Hestmann Vinjerui, Kjartan Sarheim Anthun, Andreas Asheim, Fredrik Carlsen, Bente Prytz Mjølstad, Sara Marie Nilsen, Kristine Pape, Johan Håkon Bjørngaard

## Abstract

**Introduction:** Continuous general practitioner (GP) and patient relations associate with positive health outcomes. Termination of GP-practice is unavoidable, while consequences of final breaks in relations are less explored. We will study how an ended GP-relation affects patient’s health care utilization and mortality, compared to patients with a continuous GP-relation.

**Methods and analysis:** Linking personal-level national registries data on GP-affiliation, sociodemographic characteristics, health care use and mortality, from 2008 to 2021, we will identify patients of GPs terminating practice, and match these GP-patient pairs on age, sex (both), immigrant status and education (patient), and number of patients and practice period (GPs), to GP-patient pairs with a continuous relationship in other municipalities, and compare acute and elective primary and specialist health care use mortality, before and after ended GP-practice, using Poisson regression with high-dimensional fixed effects. We will address expected modifying factors, such as patients with complex health care needs, quality of termination and GP-availability in the municipalities.

**Ethics and dissemination:** This study protocol is part of the approved project Improved Decisions with Causal Inference in Health Services Research, 2016/2158/REK Midt (the Regional Committees for Medical and Health Research Ethics) and does not require consent. We will publish in peer-reviewed journals, accessible in NTNU Open, and present at scientific conferences. To reach a broader audience, we will summarize articles in the project’s web page, regular and social media, and disseminate to relevant stakeholders.

**Strengths and limitations of this study:** This study adds to research on GP-patient relation continuity and associations, exploring termination of relations and designing a natural experiment in register data, facilitating causal inference.

The study includes the entire Norwegian population and their general practitioners (GPs) from 2008 to 2021, linking several mandatory, high-quality, healthcare, and demographic registers on a personal level.

The register data include exact time points (xxx or months? xxx) for the intervention, termination of GP-relation, and the outcomes, health care utilization and death.

By matching GP-patient pairs in practice period and distinct municipalities, we minimize time-varying confounders and dilution of effects.

As in all observational studies, our results may be influenced by residual confounding

## Background/need for research

In Norway, health care services are largely a public responsibility, where specialized health care services are provided by the state and primary care by the municipalities. General practitioners (GPs) are fundamental providers of primary care and are gatekeepers for the specialist services, which overall requires referral.

In 2001, Norway initiated the Regular GP Scheme. Under this scheme, every GP has a contract with a municipality that specifies the maximum number of patients that GP can serve, called a “patient list”. People choose their regular GP, may change GP twice a year, and will be assigned to the patient list of their preferred GP in their home municipality, provided the list is not complete. On the other hand, when a GP ends their contract with a municipality, their list patients will be randomly assigned to other GPs with contracts and capacity in the same municipality.

The regular GP Scheme aimed to assure good and equal medical care, anchored the role of gatekeeping, and the value of continuity in primary care. The scheme facilitates interpersonal continuity, an ongoing relationship between a patient and one physician, responsible for the patient’s overall health care(1). However, most GPs take part in mandatory, out-of-office public medical tasks, including out-of-hours services, resulting in absence from regular practice. In case patients require medical care when their regular GP is away, informational and “one medical home” continuity(1), is normally maintained, since most GPs work in group practices sharing electronic journal system, support staff and location and serve each other’s patients as required.

In addition to being a core value in primary care, research on GP-patient relation continuity, show associations to positive health outcomes, such as lower mortality(2) and lower specialist health care utilization(3–6). However, personal continuity is associated with practice and GP characteristics, such as number of GPs(7), list size(7,8) and perception of continuity(7), as well as patients age(7,8), sex(7,8), conditions(7,8) and number of contacts(7). These factors may also affect health outcomes, thus confounding the relationship of GP-patient continuity to health outcomes.

Breaks in continuity is less explored, but impact patients emotional stress(9). In Norway, sudden, short-term breaks showed modest changes in health care utilization, with overall less regular GP and increased out-of-hours contacts during the break, and a prolonged increase in hospital admissions for ambulatory care sensitive conditions among elderly(10).

The Regular GP Scheme has been popular, with less than 0,5% of the population opting out (11). However, the workload for GPs has increased over time, considered partly causing more GPs quitting than recruited. The GP Scheme is currently severely challenged, as one third of the municipalities do not have any available list capacity and in October 2022, more than 200 000 people lack a regular GP (3.9 % of the total population) (12). Patient ombudsmen across the country reports patients fearing lack of follow up(13) and missing a regular GP is shown to threaten basic informational continuity (14). This amplifies the need for more knowledge on health outcomes related to the definite loss of a regular GP in Norway.

We will add to former studies on GP-patient continuity, using events where original regular GPs end their contract, prompting an ended relation that is not patient-driven, aiming to avoid confounding of continuity. We will use a difference-in-difference design to compare health outcomes and health care use among the patients whose GP-relation ended to patients with a continuous GP-relation. We will further explore these effects for characteristics of the patients, the transition, and the municipalities, which may impact the outcomes, such as for patients with assumed complex needs(15), an abrupt versus gradual ended relation, and between municipalities with varying GP coverage.

## Methods

### Study period and population

In a planned ending of a GPs contract with a municipality and stopping of GP practice, we assume both GPs and patients may adjust to this, a certain time beforehand. GPs may lower their consultation activity and share the patient lists with another GP. Patients may change GP before final ending of practice. Therefore, the study period will start some years before the event that a GP ends their practice, based on an investigation of changes in practice and GP-patient continuity over approximately two to four years prior to ending. Patients will be followed for as long as available registry data permits.

The intervention population is all persons registered as list patients of GPs stopping practicing, thus being exposed to an ended GP-patient relationship. Some people experienced an ended GP-relation several times, and we studied the first episode only.

The non-intervention population is all persons registered as list patients of GPs that did not end their practice, thus exposed to a continuous GP-patient relationship, in the same study period. Some people in the non-intervention population, experienced an ended GP-relation at another time point, classifying them as intervention population and investigated as such, but not within the same study period that they were identified as part of the non-intervention population.

We matched intervention and non-intervention patients on age, sex, education, and immigrant background. Age effects on health vary with chronological age and in matching on age, we facilitate comparing patients with expected similar age effects on health over the study period.

We matched stopping and not-stopping GPs on an age span (from five years younger to five years older if the stopping GP moved, and ten years younger to equal age if the stopping GP retired), sex, and patient list length. The GPs are matched on age and practice period, to avoid bias of time trends in medical education and practice, as well as overall societal trends. This also accounts for seasonal variation. We required GPs not to practice in the same municipality, attempting to avoid spill-over effects and to facilitate comparison between municipalities.

### Data sources and variables

All citizens in Norway have a personal ID and all authorized medical doctors have a medical ID, which makes it possible to link information from the registers described below.

Statistics Norway (2006-2021) provided information on and patients birth year, sex, date for migration or death, as well as patients educational level and immigrant and country background.

The Norwegian GP Register (2006-2021) holds information on patients GP-affiliation and duration, and the GPs age, sex, specialist status, years of practice, as well as list length, vacant patient places on lists, start and end date, and cause of ended contract. The GP Register also gives information on vacant list contracts and vacant list places in the municipality overall.

The Control and Payment of Health Reimbursement register (KUHR, 2006-2021) provides information on patients’ ICPC-2 diagnoses, regular and out-of-hours primary health care utilization and identifies the GP consulted.

The Norwegian Patient Register (2008-2021) holds information on patients’ ICD-10 diagnoses, specialist health care utilization (elective and acute, outpatient visits and hospital stays, date of admission and discharge).

The Causes of Death Registry (2006-2021) gives information on patients’ date and ICD-10 diagnosed cause of death.

### Outcome variables

The main outcomes will be ordinary and out-of-hours primary care visits; elective and acute, outpatient visits and hospitalization in specialist health care; duration of hospital stays, and cause-specific mortality. We will use cause-specific outcomes for particular patient groups, such as hospital stays for ambulatory care sensitive conditions (ACSC), conditions for which hospitalization is considered avoidable with proper preventive and early disease management(16).

### Statistical analysis

We will describe sociodemographic characteristics of patients, GPs, and municipalities. We will describe GP-patient continuity for patients by having an appointed, named GP and its duration, as well as cumulative contact with the regular and other GPs. We will describe changes in practice among GPs prior to ending their contract, and for municipalities, describe GP availability at the start of the study period, as well as overall changes during 2006 to 2021.

Between 2009 and 2021, approximately 5000 GPs ended their practice, affecting closer to 5 000 000 list patients and these pairs will be matched to at least one GP-patient pair not exposed to a break in relation.

In the main analyses, we plan to use a difference-in-difference design and Poisson regression with high-dimensional fixed effects, to compare health outcomes and health care use over time among the patients whose GP-relation ended to patients with a continuous GP-relation. We will assess requirements and assumptions for the methods (pre-parallel trends and balance tests based on patient characteristics and prior health care use). In sub-studies, we will particularly explore the impact on patients with expected complex health care needs based on conditions, social and individual factors. For those with an ended relationship, we will investigate the impact of an abrupt vs gradual ended practice; the impact of varying GP coverage between municipalities; and any time trends over the complete study period.

Register data was updated including information from 2021, in January 2023. Data will be available for analysis in February/March 2023.

## Patient and Public Involvement statement

This study is part of the project Improved Decisions with Causal Inference in Health Services Research, which collaborates broadly with patient organizations, clinicians in primary and specialist health care, and the Central Norway Regional Health Authority, aiming to gain relevant input setting the overall research agenda. The steering group includes patient representatives and meets twice a year, discussing ongoing and planned studies, research questions and outcome measures, interpretation, and dissemination of results. The research group further present ongoing and planned studies in a yearly seminar open to the public.

In this study, expertise from general practitioners and primary care researchers guided the design of the study and outcome measures, and the steering group has evaluated it of great societal importance (January 2023).

## Data Availability

The data is not publicly available, but may be obtained from third parties, upon ethical approval and application.

## Abbreviations

ACPC: Ambulatory care sensitive conditions
GP: General practitioner
ICD-10: International Classification of Disease 10^th^ edition
ICPC-2: International Classification of Primary Care 2^nd^ edition

## Ethical considerations and approval

The personal data is anonymous to the researchers, and it is not possible to identify individual patients or professionals from the results. Patient consent is not required in this registry study. The Regional Committees for Medical and Health Research Ethics has approved the study, 2016/2158/REK Midt.

## Dissemination

We will publish the studies as scientific papers in peer reviewed journals and disseminate our findings at scientific conferences. Further, the regular GP scheme is of broad public interest and the findings are likely to be of interest to administrators and policy makers, as well as clinicians in both primary and specialist health care, and other researchers. We aim to present our work to all stakeholders, adapting content and form to the target group.

## Amendments

### Author statement

KHV is the project manager and drafted the protocol manuscript, while KSA, AA, JHB, FC, BPM, SMN, KP reviewed and revised it critically for important intellectual content and approved the final version for submission. All authors contributed to the research questions and study design. JHB is responsible for data acquisition, and KSA and AA prepared the data for analysis. KHV will conduct the statistical analyses. All authors will interpret the data and review, rewrite and approve study articles, while KHV will write the first drafts.

## Data statement

The data is not publicly available, but may be obtained from third parties, upon ethical approval and application. Please, contact corresponding author for statistical codes.

